# Post-Spinal Mobilization Conditioned Pain Modulation Stratifies Recovery Trajectories in Chronic Neck Pain: Toward a Mechanism-Informed Biomarker for Precision Rehabilitation

**DOI:** 10.1101/2025.09.25.25332983

**Authors:** Emmanuel Yung, Andrew Hooyman, Jo Armour Smith, Michael Wong, Jordan Cossin, Stacey Yates, Tyler Siedentopp, Shawn Farrokhi

## Abstract

**Study Design:** Prospective exploratory mechanistic cohort study.

**Objective:** To examine whether subgrouping post-spinal mobilization (post-SM) conditioned pain modulation (CPM) using grouping based on a median threshold predicts 4-week recovery and discharge outcomes better than continuous post-SM CPM values in adults with chronic mild mechanical neck pain.

**Background:** The identification of early responders to spinal mobilization remains suboptimal. Baseline CPM subgrouping has been shown to possess predictive value in pharmacology research; however, the value of post-SM CPM as an active mechanism-based biomarker remains poorly explored.

**Methods:** Thirty-one participants (18 women) with chronic neck pain received 10 minutes of cervical spinal mobilization. The post-treatment CPM responses were categorized using a median threshold to form subgroups. Associations with pain, disability, and global recovery were assessed, as were discharge rates. Receiver operating characteristic (ROC) analyses were used to identify thresholds for discharge prediction.

**Results:** Post-SM CPM subgrouping accurately predicted 4-week perceived recovery (β=0.66; 95% CI: 0.03–1.29; p=0.04), explaining >50% of the variance after adjustment for age, sex, and BMI. Likewise, the lower CPM subgroup demonstrated clinically relevant improvements in pain and function with moderate-to-large effect sizes. Discharge was also more common in this subgroup (56% vs. 13%). ROC-based thresholds optimized sensitivity for predicting discharge, but with decreasing specificity over time. However, continuous post-mobilization CPM values did not predict improvement in pain or disability scores and highlighted the exploratory nature of such results.

**Conclusions:** Post-SM CPM subgrouping can identify responders to spinal mobilization interventions. Post-mobilization stratification based on a mechanism can guide earlier discharge planning and prevent overtreatment. The quantification of CPM immediately after spinal mobilization can help clinicians guide treatment and allocate appropriate care pathways. These results must be used with caution since the sample size was small and must be investigated in larger samples.

## INTRODUCTION/BACKGROUND

Neck pain is one of the most disabling conditions globally, with 5-22% of patients presenting with mild symptoms initially, would eventually develop into more chronic severe disability within six months.[1–4] The condition places a substantial burden on workers and employers.[5–8] Spinal mobilization (SM) is a common, guideline-recommended treatment for chronic neck pain, but patient responses are quite variable.[9–10] In addition, the optimal dose of treatment sessions is uncertain—clinical guidelines often recommend up to six visits, even though > 50% of patients may be able to discharge after one visit as in low back pain.[11–12] Robust clinical tools to predict who will benefit most from SM based on the underlying pain mechanisms are lacking.[13–14] This requirement highlights the gap for biomarkers to guide personalized rehabilitation and avoid unnecessary treatments.

Conditioned Pain Modulation (CPM) reflects the pain inhibition pathway in the brain and is a promising biomarker. However, the predictive value of baseline CPM in rehabilitation has been inconsistent, highlighting the need to explore post-treatment CPM measures.[15–16] SM has been shown to change CPM responses early after treatment,[16–18] offering a dynamic method of characterizing treatment responders in real time. To date, there has been limited research on whether subgrouping patients according to their CPM response after SM can more accurately predict recovery outcomes, which is what the current study aims to accomplish.[19] Building on Yarnitsky’s work with baseline CPM subgrouping as a predictor, we used median-split subgrouping to post-treatment CPM, with a pliable and intervention-responsive indicator that can more validly forecast who recovers most optimally. This study also used receiver operating characteristic (ROC) analyses to determine optimal CPM subgroup cutoff points, which will enhance clinical prediction accuracy and advance CPM stratification in clinical rehabilitation practice. While most previous studies have focused on baseline biomarkers to predict recovery, our study introduces a dynamic, post-intervention biomarker stratification approach. This relatively uncommon framework captures treatment-responsive mechanisms in real time and may provide a more nuanced prognostic indication of perceived recovery in chronic neck pain.

## OBJECTIVES

In this study, we examined whether assessing CPM directly following a single bout of SM could categorize adults with mild mechanical chronic neck pain into distinct subgroups, whether subgroup membership was associated with 4-week outcomes, and whether pain, disability, self-perceived recovery, and discharge rates between subgroups differed over time. We hypothesized that adults with mild chronic mechanical neck pain stratified into a lower post-mobilization CPM subgroup—based on grouping by a median threshold—would exhibit greater improvements in pain, function, and perceived recovery over 4 weeks compared to those in the higher CPM subgroup. This stratification was expected to serve as a valuable early, mechanism-informed clinical indicator of treatment response.

## DESIGN

This prospective exploratory mechanistic cohort study examined the longitudinal relationships between post-spinal mobilization (post-SM) CPM (or its subgrouping) and outcomes such as pain, disability, and perceived recovery following SM in adults with mechanical chronic neck pain.

This section details the blinded protocol for measuring post-spinal mobilization CPM and applying Yarnitsky’s (2012) median-split subgrouping method post-intervention to identify mechanistic subtypes relevant for individualized clinical care.[19]

## METHODS

### Setting

The study was conducted in two academic physical therapy departments between 6/21/21 and 8/11/22. The study protocol was reviewed and approved as exempt by the Institutional Review Boards of both participating universities (IRB #2021-145). All the participants provided written informed consent in accordance with the Declaration of Helsinki. The study protocol and analysis plans were retrospectively registered on the Open Science Framework (OSF) https://doi.org/10.17605/OSF.IO/B4FME after data collection owing to evolving mechanistic hypotheses.

### Subject Recruitment

Participants were recruited between 18 and 55 years of age with chronic mild neck pain through community emails, clinic posters, and social media. Eligibility screening included exclusion of patients with a recent history of neck treatment or contraindications to spinal mobilization.[11, 20–21] The baseline Numeric Pain Rating Scale (NPRS) score was 3 or higher on a pain scale rated out of 10 (0/10= no pain, 10/10= worst pain). Their baseline Neck Disability Index (NDI) was 10%-40% out of 100% (NDI was from 0-50 divided by 50 × 100%). Additional exclusion criteria included the inability to communicate in English. Eligible participants met a research assistant who provided a detailed explanation of the study protocol and provided written informed consent. Eligible participants were instructed to refrain from taking analgesics, serotonin-norepinephrine reuptake inhibitors (SNRIs), and opioids for 24 hours before data collection.

### Demographic and Clinical Variables

Demographic and clinical variables were collected at the initial visit, including age, sex, body mass index (BMI), and use of pain medications (analgesics, serotonin and norepinephrine reuptake inhibitors, and opioids). The dependent variables were pain, measured using the Numeric Pain Rating Scale (NPRS), and function, and perceived recovery, were measured using the Neck Disability Index (NDI) and Global Perceived Recovery (GPR), respectively. We interpreted the GPR rating recommended by Cote et al. (2016) as valid and reliable as follows: (7) worse than ever, (6) much worse, (5) slightly worse, (4) no change, (3) slightly improved, (2) much improved, and (1) completely better. Patients whose responses were either (1) or (2) indicated that they recovered and were ready for discharge when this measure was used in the clinical setting.[11]

### Conditioned Pain Modulation/Spinal Mobilization Procedures

A fellowship-trained, board-certified orthopaedic clinical specialist physical therapist with over 20 years of experience performed a clinical examination to assess the active range of motion of the patient’s cervical spine. A manual examination was also conducted to identify the most painful segmental level using both the unilateral posterior-anterior (PA) and anterior-posterior (AP) oscillatory techniques. This was the same segmental level at which PA and AP were applied. A 15-minute rest period was followed, during which the investigator provided information on the natural history of neck pain, how SM is believed to work, and the ability to fully participate in usual activities after the intervention. The participants were also instructed to avoid other SM or exercise treatments during the 4-week follow-up period.

Conditioned Pain Modulation (CPM) was assessed by measuring pain pressure thresholds (PPTs) at three anatomical sites before (PPT1) and after (PPT2) a cold pressor test and immediately after spinal mobilization (PPT3 and PPT4). The detailed measurements are presented in Table 3.[22–24] CPM is calculated as follows, where a positive number signifies a normal CPM and no change or a negative number indicates an abnormal or impaired CPM, and is expressed as a %.

After completing the baseline CPM assessment, participants received 10 minutes of grade III unilateral PA and AP oscillatory spinal mobilization, delivered in a randomized order to the most symptomatic cervical segment.

This SM dose was based on a study by Lascurain-Aguirrebena (2019), who demonstrated improved pain and perceived global effects.[25] The same investigator provided SM to all participants and was blinded to all baseline and follow-up measurements collected by a research assistant. In addition, a second licensed physical therapist, not involved in treatment or assessment, randomly observed sessions to verify adherence to blinding and protocol fidelity.[26] These measures and the randomization of the order of CPM testing and SM delivery reduced the likelihood of bias.

Immediately following the SM intervention, post-mobilization CPM was measured using the same procedure as that used for baseline conditioned pain modulation. For the post-mobilization CPM measures, PPT1 was renamed PPT3 and PPT2 was renamed PPT4. The change in conditioned pain modulation was calculated as the difference between post-mobilization CPM and baseline CPM. Following completion of the in-person portion of the study, follow-up emails with instructions for participants to complete the NPRS, NDI, and GPR were sent out two days post, 2-weeks post, and 4-weeks post-SM. Non-responders received follow-up phone calls to maximize data completeness, and participants were instructed to avoid any additional spinal mobilization or exercise treatment during the 4-week follow-up period to minimize confounding factors.

### Statistical Analyses

#### Baseline Characteristics

Participant characteristics, including demographics and baseline clinical variables, were first summarized using descriptive statistics for continuous variables (e.g., age, NPRS, GPR, NDI, and BMI) and percentages (counts) for categorical variables (e.g., sex).

#### Longitudinal Analyses

Separate linear mixed-effect models were used to assess whether post-mobilization CPM was associated with a longitudinal trend in change in pain (NPR), function (NDI), and GPR. The primary predictive variable was post-mobilization CPM. The variable time was treated as a continuous variable with: Baseline (day 0), followed by 2-day, 2-week, and 4-week follow-up. Each model was adjusted for baseline patient demographics (age, sex, and BMI). Individual participant identifiers were used to model random intercepts within the model.

#### Exploratory subgroup analysis for prediction of clinical outcomes and discharge

Researchers have dichotomized CPM as a predictor of the selective efficacy of SNRIs, as they would appear ineffective when the cohort is not subgrouped.[19] Similarly, we used a mixed-effect ANOVA to explore whether the dichotomized post-mobilization CPM subgroups were associated with the 4-week outcomes (e.g., NPRS, NDI, and GPR). Dichotomizing the post-mobilization CPM using grouping based on a median threshold placed individuals into more-versus less-efficient pain control subgroups. Therefore, separate models were conducted for each clinical outcome as the dependent variable, and time, CPM subgroup, and other covariates were included as fixed effects, with random intercepts on participants. In addition to modeling clinical outcomes, analyses to examine the relationship between participant discharge and post-mobilization CPM were also performed. Separate logistic regression models were generated to predict discharge at the various follow-up periods (2-day, 2-week, 4-week). Importantly, global perceived recovery (GPR) at baseline is based upon the participants’ expectation of whether or not they will be discharged by 4-week, corresponding to GPR of 1 and 2 (discharged) versus 3 through 7, respectively. Discharge at 2-days, 2-week and 4-week are designated as true discharge based on GPR scores of 1 or 2. Model metrics of sensitivity, specificity, positive predictive value, negative predictive value, and area under the curve, were computed to evaluate its clinical utility. Optimal cut-points for post-mobilization CPM for each time point were calculated using Youden’s index. The resulting cut-points were then used to calculate differences in 4-week GPR, NDI, and NPRS between predicted discharge and non-discharge patients. These results were then compared to a simple median split of post-mobilization CPM to compare how identification of optimal cut-points based on discharge varies clinical outcomes. All statistical analyses were performed using R 4.0.3.

## RESULTS

From an initial total of 151 respondents to the flier and email advertisements, 31 participants (18 women) completed the study, including all 3 follow-up periods. Fifty of the 151 respondents did not respond when the research assistants attempted to schedule appointments, whereas 61 were deemed ineligible. Two people canceled their appointments, six were ineligible following in-person screening/evaluation, and one completed in-person data collection and was excluded because of a lack of follow-up (Figure 1). Figure 1 shows the study flow and mechanistic framework explored in this study, including baseline CPM measurement, spinal mobilization, post-intervention CPM measurement, and median-split subgrouping. These groups were associated with differences in recovery at four weeks but not with pain, disability, or discharge rate outcomes.

**Figure 1.**
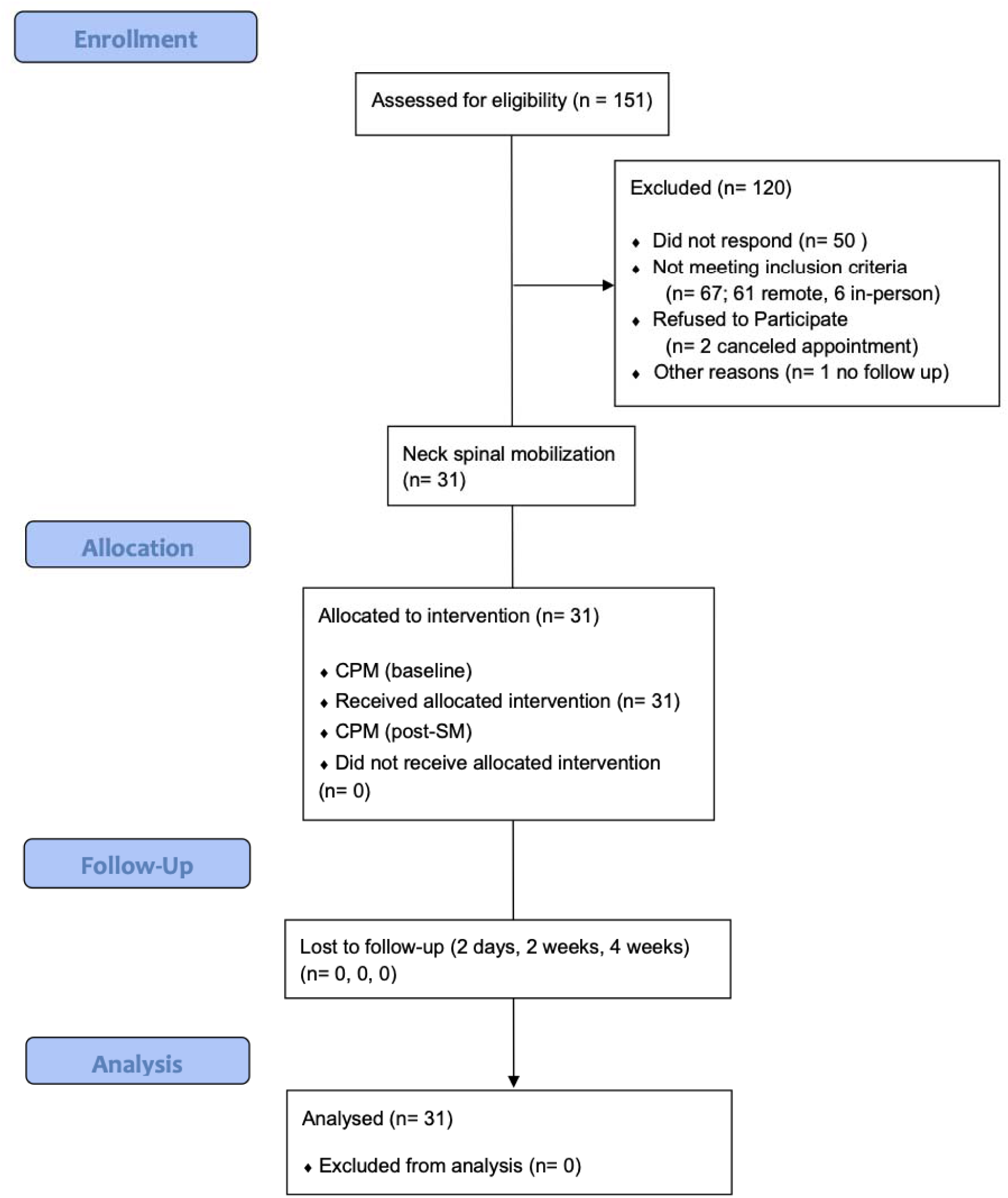
Study flow and mechanistic framework.

Figure 1 shows study flow (above) and mechanistic framework (here): Baseline conditioned pain modulation (CPM), spinal mobilization (SM), post-spinal mobilization (post-SM) CPM measurement, median-split subgrouping, and their associations with 4-week perceived recovery. No significant association with pain, disability, or discharge was observed.

Table 1 presents participants’ demographic, anthropometric, and clinical outcomes from baseline to follow-up. The discharge count was the total number of participants who reported or were discharged during the follow-up period.

**Table 1.**
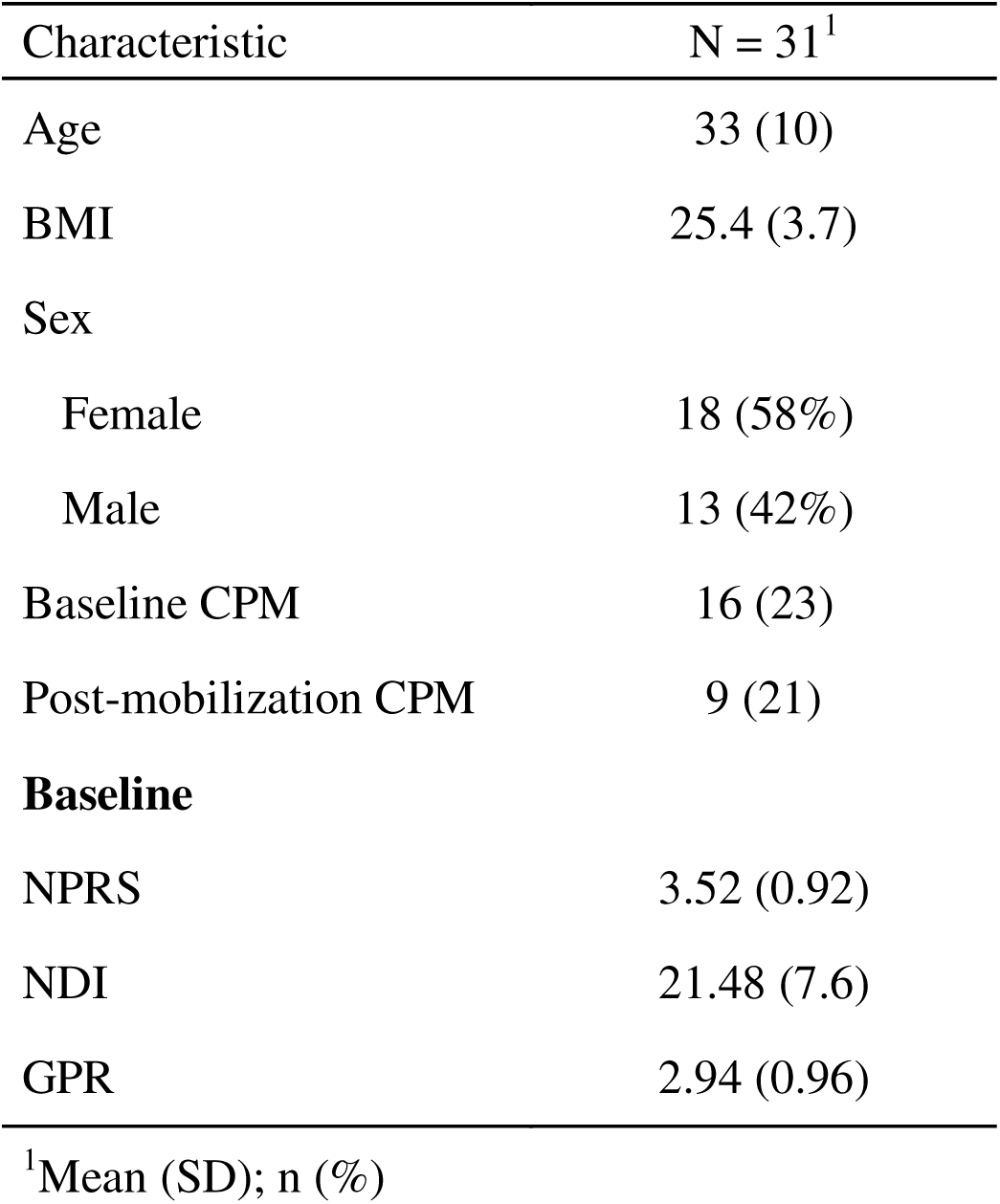
Participant characteristics

| Characteristic | N = 31<sup>1</sup> |
| --- | --- |
| Age | 33 (10) |
| BMI | 25.4 (3.7) |
| Sex |  |
| Female | 18 (58%) |
| Male | 13 (42%) |
| Baseline CPM | 16 (23) |
| Post-mobilization CPM | 9 (21) |
| **Baseline** |  |
| NPRS | 3.52 (0.92) |
| NDI | 21.48 (7.6) |
| GPR | 2.94 (0.96) |

<sup>1</sup>Mean (SD); n (%)

Body mass index (BMI), Conditioned Pain Modulation (CPM), Numeric Pain Rating Scale (NPRS) score, function, and perceived recovery were measured using the Neck Disability Index (NDI) and Global Perceived Recovery (GPR).

### Longitudinal Association between post-Mobilization CPM and Clinical Outcomes

Longitudinal analyses revealed that continuous post-mobilization CPM values were not predictive of changes ly in pain or disability. However, median split subgrouping revealed distinct recovery trajectories and discharge differences (Tables 2 and 3). Detailed model estimates, including the effect sizes and interactions, are presented in these tables.

**Table 2:** Full mixed-effects model estimates for GPR(Table 2A), NDI (Table 2B), NPRS (Table 2C), including p-values and confidence intervals.

| GPR |  |  |  |  |
| --- | --- | --- | --- | --- |
|  |  | Lower | Upper |  |
|  |  | 95% | 95% |  |
|  | Estimate | CI | CI | p-value |
| Intercept | 2.696 | 0.575 | 4.817 | *0.013 |
| Time (day) | 0.014 | 0.001 | 0.026 | *0.034 |
| Post-Mobilization |  |  |  |  |
| CPM (scaled) | 0.113 | -0.228 | 0.454 | 0.502 |
| Age | 0.003 | -0.028 | 0.034 | 0.835 |
| BMI | 0.001 | -0.088 | 0.09 | 0.981 |
| Sex (Male) | 0.013 | -0.627 | 0.653 | 0.968 |
| TIME: Post- |  |  |  |  |
| Mobilization CPM |  |  |  |  |
| (scaled) | 0.013 | 0.001 | 0.026 | *0.038 |

NDI
|  |  | Lower | Upper |  |
| --- | --- | --- | --- | --- |
|  |  | 95% | 95% |  |
|  | Estimate | CI | CI | p-value |
| Intercept | 25.047 | 9.391 | 40.703 | *0.002 |
|  |  |  |  | *<0.00 |
| Time (day) | -0.235 | -0.341 | -0.129 | 1 |
| Post-Mobilization |  |  |  |  |
| CPM (scaled) | -0.321 | -2.904 | 2.261 | 0.8 |
| Age | 0.1 | -0.13 | 0.33 | 0.378 |
| BMI | -0.425 | -1.084 | 0.235 | 0.197 |
| Sex (Male) | 0.449 | -4.271 | 5.17 | 0.846 |
| TIME: Post- |  |  |  |  |
| Mobilization CPM |  |  |  |  |
| (scaled) | 0.108 | 0.002 | 0.215 | *0.047 |

NPRS
|  |  | Lower | Upper |  |
| --- | --- | --- | --- | --- |
|  |  | 95% | 95% |  |
|  | Estimate | CI | CI | p-value |
| Intercept | 4.17 | 1.603 | 6.737 | *<0.00 |
|  |  |  |  | 1 |
|  |  |  |  | *<0.00 |
| Time (day) | -0.024 | -0.041 | -0.008 | 1 |
| Post-Mobilization |  |  |  |  |
| CPM (scaled) | 0.104 | -0.316 | 0.524 | 0.61 |
| Age | 0.04 | 0.002 | 0.078 | *0.04 |
| BMI | -0.1 | -0.208 | 0.008 | 0.07 |
| Sex (Male) | 0.093 | -0.681 | 0.867 | 0.81 |
| TIME: Post- |  |  |  |  |
| Mobilization CPM |  |  |  |  |
| (scaled) | 0.012 | -0.005 | 0.029 | 0.15 |

**Table 3.** Post-spinal mobilization (Post-SM) CPM median split subgroup comparison.

| | Post-SM CPM $\leq 6.75$ | | p-value | Cohen's d |
| --- | --- | --- | --- | --- |
|  | Yes (n=16) | No (n=15) |  |  |
|  | Mean (SD) | Mean (SD) |  |  |
| Age | 33 (med) | 27 | 0.21 | n/a |
| BMI | 25.53 | 25.27 | 0.85 | n/a |
| Sex | 0.50 males | 0.33 males | 0.34 | n/a |
| (Baseline) PPT1 | 17.66 (8.1) | 12.53 (5.4) | **0.047\*\*** | **0.74 (e)** |
| (Post Cold Pressor Test) | 18.9 (8.9) | 15.1 (5.9) *ppt1 vs | 0.28 | n/a |
| PPT2 |  | 2 |  |  |
| (Post SM) PPT3 | 19.72 (7.9) | 13.84 (6.2) | **0.02\*\*** | **0.82 (f)** |
| (Post SM + Cold Pressor) |  | 17.24 (7.9) *ppt3 |  |  |
| PPT4 | 18.49 (8.3) | vs 4 | 0.68 | n/a |
| Baseline CPM | 8.17 (19.0) | 24.98 (24.6) | **0.04\*\*** | **0.75 (e)** |
| Post-SM CPM | -6.28 (13.2) | 24.55 (15.6) | **0.00\*\*** | **2.08 (f)** |
| CPM change | **-14.4 (19.0)\* (e)** | -0.43 (26.9) | **0.046\*\*** | **0.59 (e)** |
| Baseline NPRS | 3.69 (1.1) | 3.33 (0.6) | 0.28 | n/a |
| Change at 2-days | **-1.29 (1.1)\* (c,f)** | -0.76 (0.8)* (f) | 0.13 | n/a |
| Change at 2-weeks | **-1.54 (1.6)\* (c,f)** | -0.56 (1.2) | 0.07 | n/a |
| Change at 4-weeks | **-1.63 (1.9)\* (c,e)** | -0.49 (1.4) | 0.07 | n/a |
| Baseline NDI | 22.62 (6.6) | 20.27 (8.6) | 0.40 | n/a |
| Change at 2-days | **-9.38 (7.9)\* (c,f)** | -6.13 (6.9)* (f) | 0.23 | n/a |
| Change at 2-weeks | **-9.25 (8.5)\* (c,f)** | -6.13 (9.5)* (e) | 0.35 | n/a |
| Change at 4-weeks | **-13.38 (11.2)\* (c,f)** | -5.73 (11.3)* (e) | 0.07 | n/a |
| Baseline GPR | 2.75 (1.0) | 3.13 (0.9) | 0.28 | n/a |
| Change at 2-days | -0.25 (1.2) | -0.27 (0.7) | 0.96 | n/a |
| Change at 2-weeks | 0.06 (1.0) | 0.4 (1.1) | 0.40 | n/a |
| Change at 4-weeks | -0.12 (1.3) | 0.6 (1.4) | 0.14 | n/a |
| % discharged at 4-weeks | **56% (9/16)** | 13.3% (2/15) | **0.013\*\*** | **0.45 (h)** |

Complete results of each linear mixed-effects model with estimates, 95% confidence intervals, and p-values for all fixed effects. The significant results are denoted by *. Conditioned Pain Modulation (CPM), body mass index (BMI), Numeric Pain Rating Scale (NPRS), function, and perceived recovery were measured using the Neck Disability Index (NDI) and Global Perceived Recovery (GPR).

While continuous post-SM CPM values alone did not predict outcomes, post-SM CPM stratification using grouping based on a median threshold identified subgroups with varying recovery patterns and strongly differing 4-week discharge rates, revealing the practical importance of post-intervention CPM subgrouping. This disparity in discharge rate underscores the strong clinical value of the post-SM CPM subgrouping, highlighting its potential as a simple-to-use stratification tool for guiding early treatment.

Using the median threshold of 6.75%, two subgroups emerged: 16 participants with post-SM CPM ≤6.75 and 15 above the threshold. Table 3 provides the post-mobilization CPM median split subgroup’s comparative demographics, PPT values, CPM measures, and clinical outcomes over 2-days, 2-weeks, 4-weeks. Participants with post-SM CPM ≤6.75 demonstrated significantly lower baseline CPM (mean 8.17 vs. 24.98, p=0.04), greater CPM change (−14.4% vs. -0.43%, p=0.046), and superior clinical improvements in pain and disability exceeding Minimal Clinically Important Differences (MCID) with moderate to large effect sizes. Of particular interest, 56% of the lower CPM subgroup was discharged by 4 weeks in contrast with 13% of the higher CPM subgroup (p=0.013), highlighting the clinical value of stratification at an early stage. These findings demonstrate the magnitude of clinical utility: post-SM CPM subgrouping explained > 50% of the variance in 4-week perceived recovery outcomes and pain/disability improvement often exceeded the established MCID thresholds, indicating subgroup identification as a potentially actionable clinical tool.

Between subgroup comparisons based on individuals who reporting global perceived recovery scores of 1 or 2 (representing discharge criteria when used in clinical practice) and those who did not.

Significant difference: p-value < 0.05

a. Within subgroup significant difference (compared to baseline)*
b. Between subgroup significant difference** (2-sample T-test or Mann-Whitney)
c. Exceeds MCID for NPRS (1.0)[27]; NDI (7%)[28]

(d-f) Effect size (Cohen’s d): small >0.2 (d), medium >0.5 (e), large >0.8 (f)

(g-i) Effect size (Cohen’s w): small >0.1 (g), medium >0.3 (h), large >0.5 (i)

(Parametric comparison) between group: 2 proportion test; within group: paired T-test

(Nonparametric comparison) Within group/s: Mann-Whitney; within group: 1-sample Wilcoxon

Conditioned Pain Modulation (CPM), body mass index (BMI), pain pressure thresholds (PPTs), Numeric Pain Rating Scale (NPRS), function, and perceived recovery were measured using the Neck Disability Index (NDI), Global Perceived Recovery (GPR), and minimal clinically important difference (MCID).

Table 4 results show that post-SM CPM subgrouping was significantly associated with 4-week perceived recovery, explaining over 50% of variance after adjusting for covariates. No significant predictive value was found for pain (NPRS) or disability (NDI).

**Table 4.** Longitudinal analysis using mixed effects modeling Estimated effects (β) of post-spinal mobilization (post–SM) CPM-based subgrouping adjusted for 279 time, baseline CPM, and demographics (age, sex, and BMI) on pain (NPRS), disability (NDI), 280 and recovery (GPR) over 4-weeks. CPM: conditioned pain modulation; NS: not significant

| Outcome Measure | Fixed Effect | Estimate ($\beta$) | 95% CI | p-value | R<sup>2</sup> (Adj)% | Interpretation |
| --- | --- | --- | --- | --- | --- | --- |
| Global Perceived Recovery (GPR) | Group | 0.66 | 0.03, 1.29 | 0.04 | 50.52 | Post-SM CPM-based subgrouping predicts recovery |
| Numeric Pain Rating Scale (NPRS) | Group | NS |  | >0.05 |  | No significant predictive value |
| Neck Disability Index (NDI) | Group | NS |  | >0.05 |  | No significant predictive value |

Estimated effects ( $\beta$ ) of post-spinal mobilization (post-SM) CPM-based subgrouping adjusted for time, baseline CPM, and demographics (age, sex, and BMI) on pain (NPRS), disability (NDI), and recovery (GPR) over 4-weeks. CPM: conditioned pain modulation; NS: not significant

Figure 2 illustrates individual progression of global perceived recovery (GPR) and disability (NDI) clinical outcomes across the experimental follow-up period. Individual points and lines represent individual data. The blue line represents the fitted slope of individuals with post-mobilization CPM one standard deviation below the average, the purple line represents the fitted slope of individuals with average post-mobilization CPM, and the red line represents individuals with post-mobilization CPM one standard deviation above the average. Figure 3 shows the discharge rates comparison post-spinal mobilization CPM median-split subgroups over time

**Figure 2:**
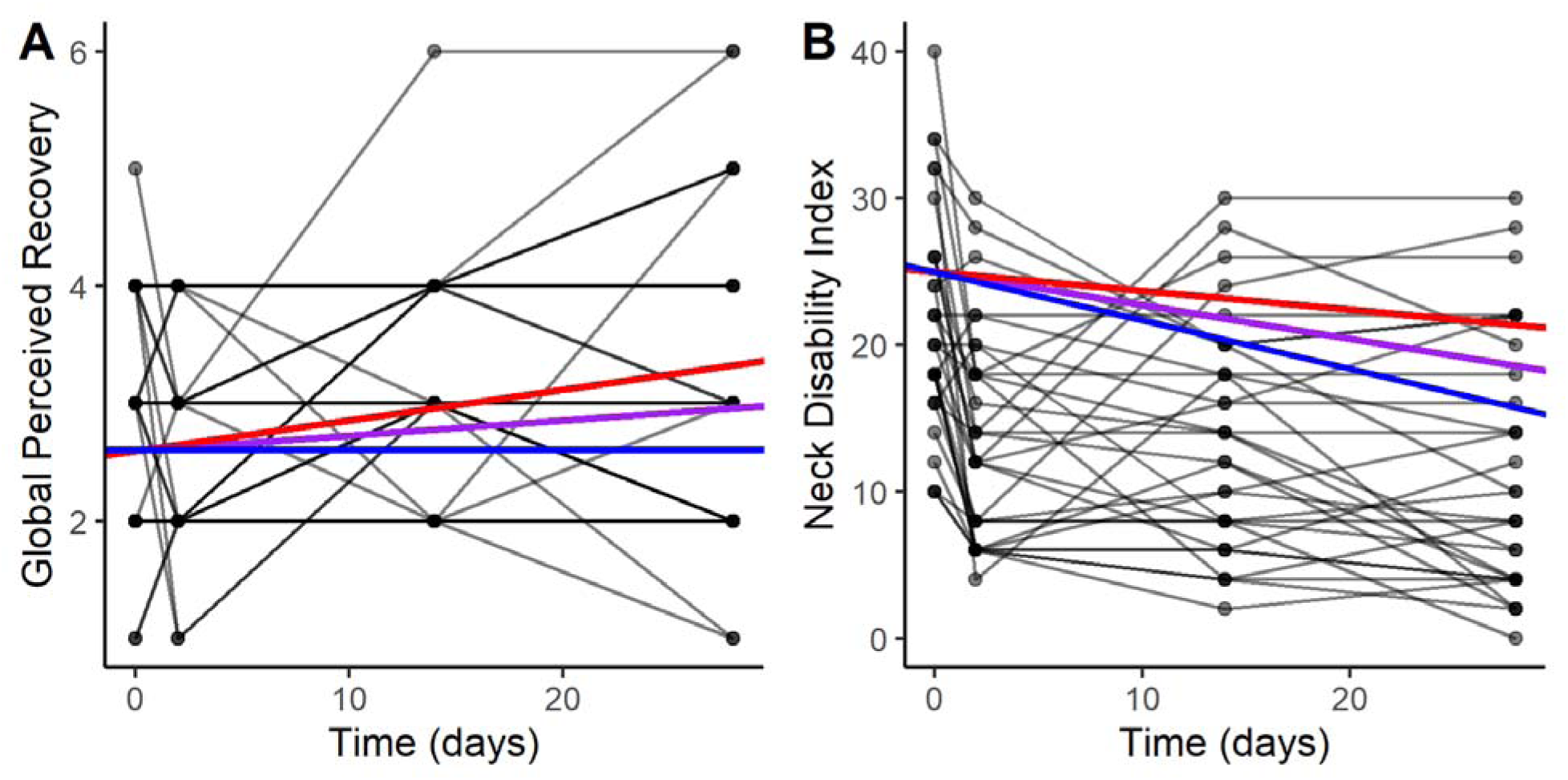
Longitudinal Visualization of Recovery Trajectories

**Figure 3.**
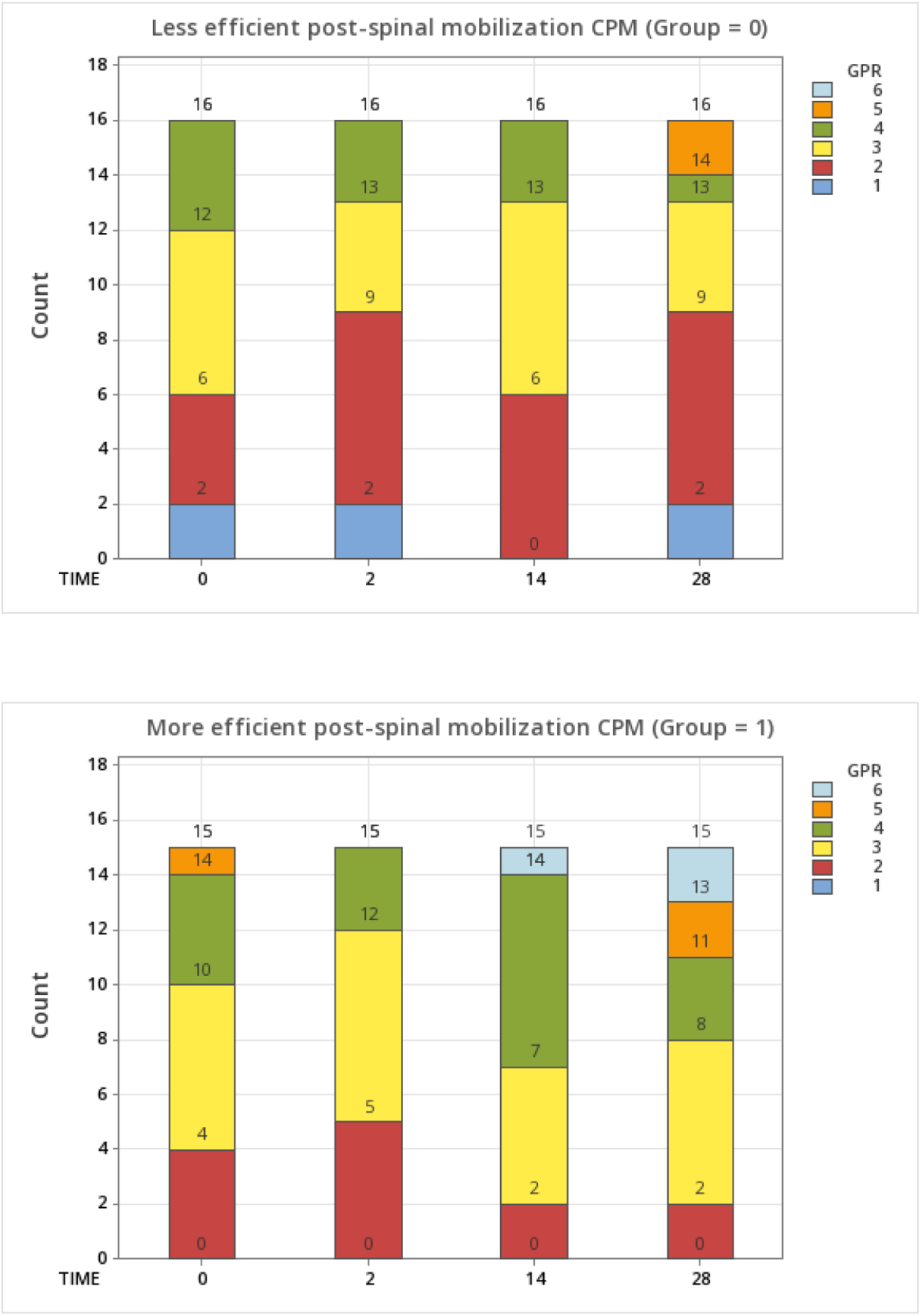
Discharge rates comparison post-spinal mobilization CPM median-split subgroups over time

Fitted mean recovery slopes ±1 SD based on continuous post-spinal mobilization (post-SM) conditioned pain modulation (CPM). This figure illustrates the differences in recovery trajectory.

Post-spinal mobilization conditioned pain modulation (CPM) was dichotomized via a median split so that: =/< 6.75 (less efficient CPM, Group 0, top figure) and > 6.75 (more efficient CPM, Group 1, bottom figure). GPR= global perceived recovery, actual GPR at three follow-up timepoints (2-day, 14-day, 28-day) with the expected 4-week GPR as reference on day 0.

### Identification of Post-Mobilization Cut Points Predictive of Discharge

The diagnostic accuracy of the post-mobilization CPM optimal cutoff points across different follow-up periods is shown in **Table 5**. At 2 days, post-mobilization CPM cut point of 13.11 demonstrated a sensitivity of 0.57 (95% CI: 0.29–0.82) and specificity of 0.82 (95% CI: 0.57– 0.96), with positive predictive value (PPV) of 0.73 (95% CI: 0.39–0.94) and negative predictive value (NPV) of 0.70 (95% CI: 0.46–0.88). The approximate area under the curve (AUC) was 0.71 (95% CI: 0.52–0.91).

**Table 5.**
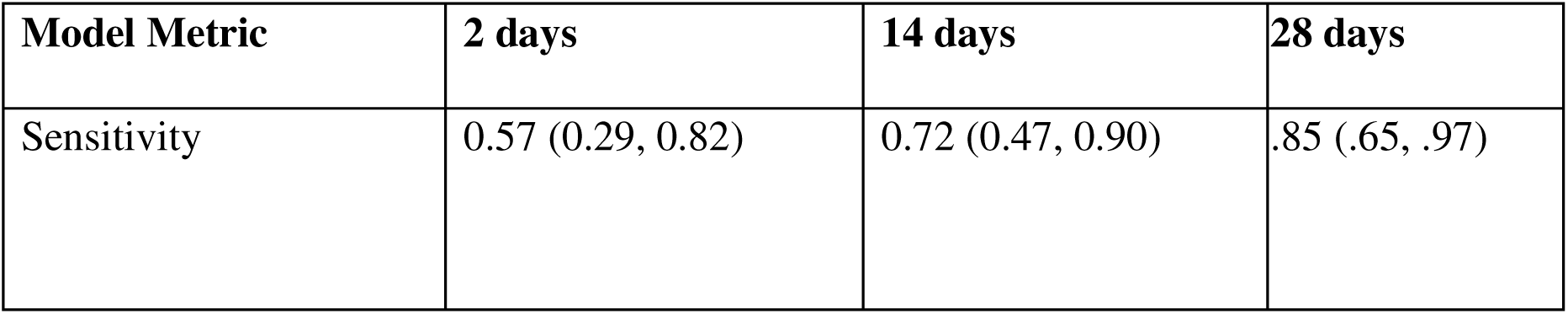
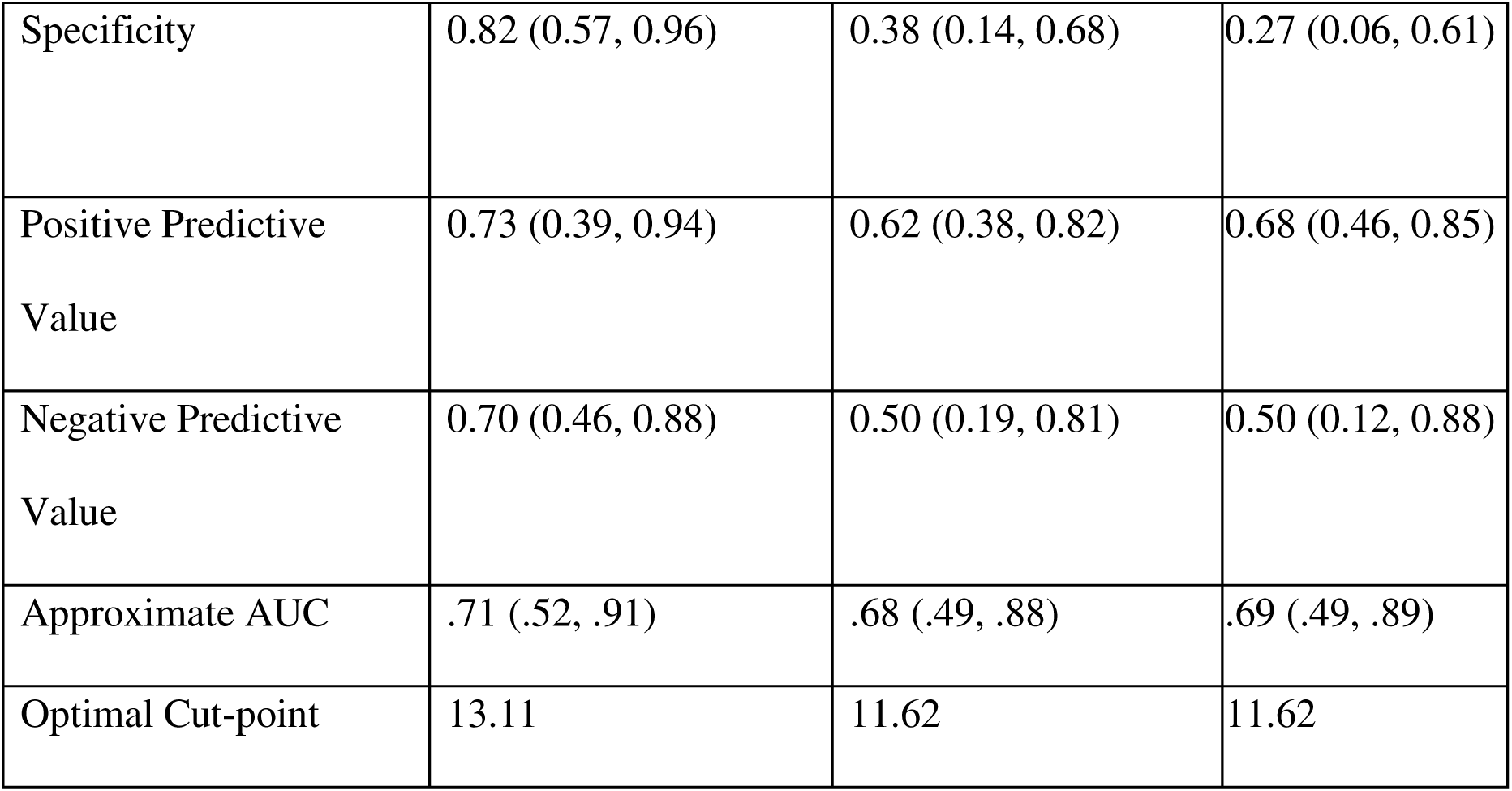
Various model metrics and outcomes for the different post-spinal mobilization Conditioned Pain Modulation models predicting discharge over time. Various model metrics and outcomes for different post-spinal mobilization Conditioned Pain Modulation (CPM) models to predict discharge on different days of the follow-up period. 95% Confidence Intervals are within parentheses. Area under the curve (AUC).

At 14 days, the post-mobilization CPM optimal cut point was 11.62 and the sensitivity improved to 0.72 (95% CI: 0.47–0.90), while specificity decreased to 0.38 (95% CI: 0.14–0.68), with PPV and NPV of 0.62 (95% CI: 0.38–0.82) and 0.50 (95% CI: 0.19–0.81), respectively. The AUC at this time point was 0.68 (95% CI: 0.49–0.88).

By 28 days, the post-mobilization CPM optimal cutoff point was also 11.62 the sensitivity further increased to 0.85 (0.65–0.97), but the specificity dropped to 0.27 (0.06–0.61). The PPV remained robust at 0.68 (0.46–0.85), while the NPV was 0.50 (0.12–0.88). AUC was 0.69 (0.49– 0.89). Overall, the model was more sensitive but less specific over the period, indicating an increased ability to identify correct discharges but with higher rates of false positives.

The visualization of each ROC curve for each discharge time (day-2, week-2, and week-4) is shown in Figure 4. Notably, optimized cutoff points enhanced predictive sensitivity compared with the median split, highlighting the importance of data-driven threshold refinement.

**Figure 4.**
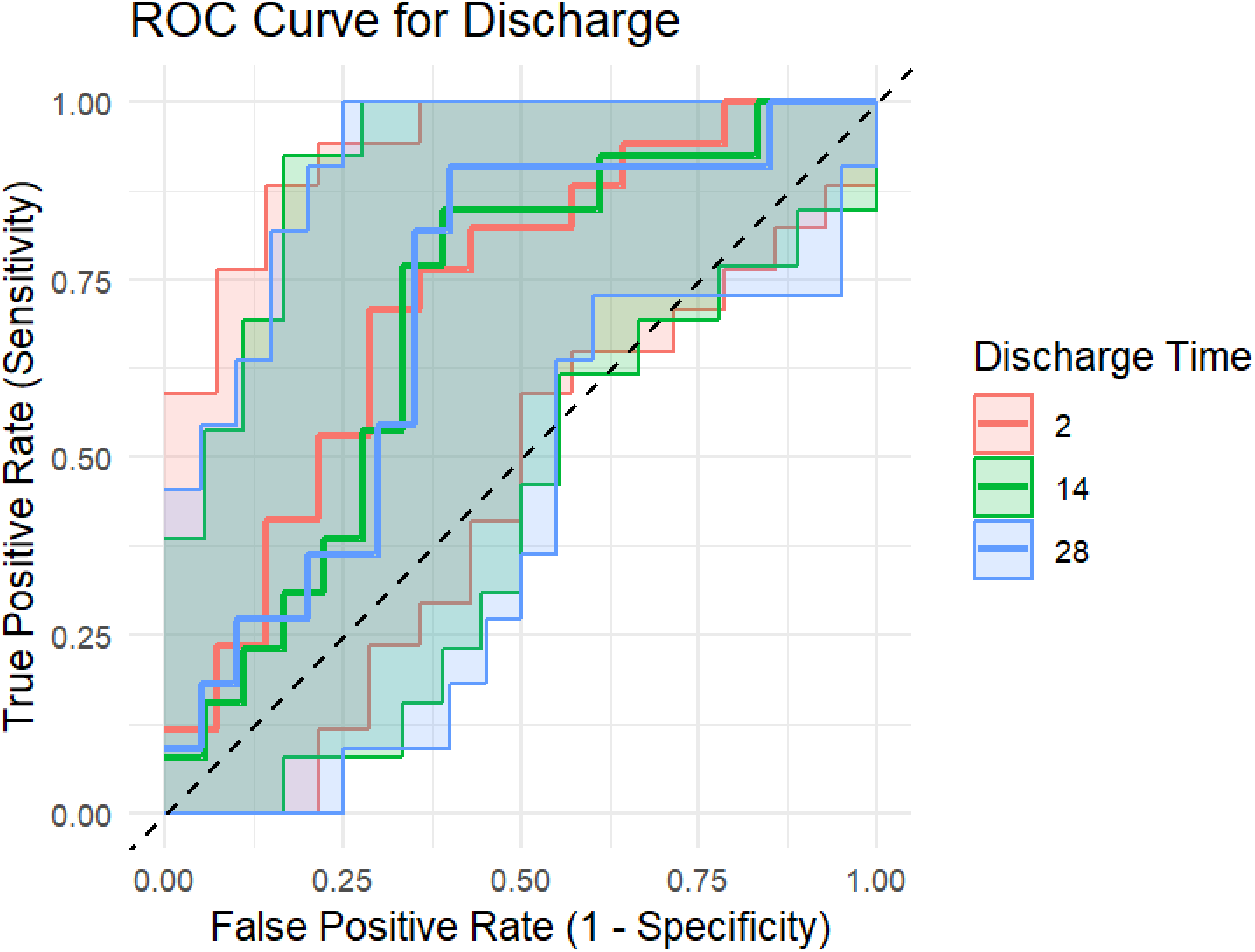
Different receiver operator characteristic curves specific to discharge rates at different follow-up times for post-mobilization CPM. The dashed line along the diagonal represents the no information criterion.

Summary clinical outcome statistics by post-mobilization CPM median split or optimal cutoff point subgroups indicated that members in the low post-mobilization CPM subgroup had an increased proportion of discharge and better clinical outcomes than those in the high post-mobilization CPM subgroup. The statistics are presented in Table 6.

**Table 6.** Clinical outcomes stratified by post-spinal mobilization CPM using both median split and optimized ROC-derived cut points. Outcomes consistently favored the lower post-mobilization CPM or “predicted discharge” subgroup across time points. ^D^Discharge count represents the total number of participants who report or are discharged from that follow-up period and onward. Conditioned Pain Modulation (CPM), Numeric Pain Rating Scale (NPRS) score, function, and perceived recovery were measured using Neck Disability Index (NDI) and Global Perceived Recovery (GPR). These findings are exploratory and should be cautiously interpreted. Further studies with larger sample sizes are required to validate our results.

| **CPM Split Optimal**<br>**Cut Point** | **$\leq 6.75$** | **$> 6.75$** | **$\leq 13.11$** | **$> 13.11$** | **$\leq 11.62$** | **$> 11.62$** |
| --- | --- | --- | --- | --- | --- | --- |
| **Split Type** | **Median** |  | **Day 2** |  | **Day 14/28** |  |
| **Patient Count** | **16** | **15** | **19** | **12** | **18** | **13** |
| **Discharge Count (n)<sup>□</sup>** | 9 | 2 | 14 | 3 | 11/10 | 2/1 |
| **NPRS (Pain) Mean ±**<br>**SD** | 2.03 ±<br>1.94 | 2.84 ±<br>1.33 | 2.19 ±<br>1.81 | 2.83 ±<br>1.48 | 2.14 ±<br>1.85 | 2.84 ±<br>1.41 |
| **NDI (Function) Mean**<br>**± SD** | 9.25 ±<br>8.88 | 14.53 ±<br>7.83 | 9.89 ±<br>8.78 | 14.83 ±<br>7.92 | 9.67 ±<br>8.98 | 14.77 ±<br>7.6 |
| **GPR (Perceived**<br>**Recovery) Mean ± SD** | 2.62 ±<br>1.2 | 3.73 ±<br>1.28 | 2.84 ±<br>1.38 | 3.67 ±<br>1.15 | 2.78 ± 1.4 | 3.69 ±<br>1.11 |

## DISCUSSION

This exploratory analysis addresses a gap highlighted in prior reviews, in which the failure to detect CPM treatment associations may stem from analyzing cohorts without stratification. By revealing responders through post-SM CPM subgrouping, this stratification offers a promising means of customizing treatment interventions, potentially reducing unnecessary care and associated costs. These findings align with personalized care and responsibly utilize healthcare resources. Importantly, the post-SM CPM subgrouping explained over 50% of the variance in the 4-week perceived recovery, which is a substantial effect of a mechanism-based biomarker. The lower CPM subgroup displayed clinically meaningful improvements; over half (56%) achieved discharge within 4 weeks versus just 13% in the higher CPM subgroup, and improvements in both pain and disability consistently exceeded the Minimal Clinically Important Differences (MCID; NPRS ≥1, NDI ≥7%), indicating tangible benefits beyond statistical significance. This immediate clinical effect underscores the translational significance of the subgroup that targeted earlier stratification. We observed differential effects on perceived recovery, suggesting that this may be an early mechanistic marker of SM.[13] This subgroup’s CPM change post-SM may have interacted with important serotonergic and noradrenergic pathways, potentially benefitting nociplastic pain.[13] Thus, subgrouping might provide a preliminary indication of SM-related potential for clinical recovery. With respect to this view, analysis of the overall cohort without stratification based on CPM subdued the potential prognostic impact of post-mobilization CPM. This highlights the advantage of a subgrouping approach in the prediction of treatment response at the individual level.[16] This was because the subgroups reacted differently to SM, with their corresponding CPM almost cancelling each other out, as in our study. CPM-threshold subgrouping was also associated with perceived recovery when applied to stratifying participants in mechanism-based analysis. Conversely, participants above the CPM threshold had little change in CPM or clinical response and perhaps lesser short-term benefit from additional SM, rendering them potential candidates for a different kind of intervention.

These findings contribute to the evolving paradigm of prognostic modeling by highlighting the possibility of post-spinal mobilization conditioned pain modulation as a dynamic biomarker. Unlike static baseline biomarkers, this approach underscores the mechanistic and clinical significance of detecting treatment-induced neuroplastic changes that may enhance stratification of patient recovery trajectories and enable precision rehabilitation. Then, the nonresponsive patient subgroup to SM may have an underlying mechanism of pain (e.g., exercise-induced hypoalgesia) that may be more responsive to other interventions, such as, exercise.[29]

To our knowledge, this is the first exploratory cohort study to investigate stratification based on the post-mobilization CPM response, which demonstrates a potentially important correlation between short-term recovery outcomes and discharge differences among individuals with mild chronic neck pain. Conceptually guided by Yarnitsky’s 2012 study, our post-intervention CPM-based subgrouping used a similar median-split approach to stratify treatment responses.[19] Yarnitsky’s (2012) seminal baseline CPM median split identified distinct pain modulation phenotypes that were predictive of pharmacological responses.[19] Our study builds on this work by utilizing the same subgrouping based on a median threshold post-intervention in terms of CPM responsiveness after spinal mobilization as a novel biomarker for clinical recovery stratification. Most importantly, contrary to conventional expectations, lower post-SM CPM values were predictive of improved recovery and discharge status. Although we did not test whether such diminished CPM values reverted to baseline over time, the post-treatment CPM changes observed may reflect adaptive recalibration in central pain modulation, which is consistent with neuroplastic changes. However, as this investigation only measured CPM at one post-intervention time point, the duration and time course of such changes are uncertain and require further longitudinal studies. This interpretation aligns with similar transient patterns reported in exercise-induced hypoalgesia and autonomic regulation literature, supporting CPM’s potential role as a *dynamic* biomarker of recovery.

Baseline CPM has shown limited value in outcome prediction, with a systematic review reporting inconsistent findings when whole cohorts were analyzed without stratification. [30] A possible explanation for this is that the CPM shifts after treatment. Post-intervention alterations, but not baseline values, may better interpret a patient’s intervention-responsive phenotype. Evidence from the synthesis of other rehabilitation interventions corroborates this perspective. Meta-analytic evidence substantiates that spinal treatments (including motor control training, neural mobilization, and TENS) can improve impaired baseline CPM in musculoskeletal conditions. [31–32] The results above indicate that subgrouping based on CPM after intervention can recognize mechanism-responsive patients more than a particular intervention. Our post hoc ROC analyses build upon the median-split approach by specifying optimized data-driven CPM thresholds that increase the discharge prediction sensitivity while maintaining acceptable specificity. This is an essential innovation that has been shown to result in significant changes in predictive accuracy and clinical utility. The sensitivity improvement over time reflects optimized discrimination of true discharge candidates, while the reduction in specificity reflects a trade-off between prognostic biomarker optimization.

Acute reductions in conditioned pain modulation (CPM) following treatment are paradoxically related to better clinical outcomes, suggesting transient neurophysiological adaptations in the descending pain modulatory pathways. Temporary reduction of CPM function is more likely to be indicative of adaptive recalibration rather than pathology and reflects a responsive, sensitive central nervous system undergoing healthy neuroplastic changes in response to manual therapy. Similar transient responses have been reported in research on exercise-induced hypoalgesia and autonomic regulation, in which the initial exacerbation phases were followed by sustained clinical improvement.[33] Such transient oscillations are indicative of dynamic recalibration in descending inhibitory pathways and identify CPM as a valuable biomarker of recovery. This multifaceted interplay between the autonomic nervous system response, central sensitization, and neuroplastic mechanisms positions post-mobilization CPM alterations within the framework of a time-limited, potentially adaptive process from an evolutionary perspective that can provide valuable information for individualized rehabilitation strategies. These interpretations remain speculative and should be tested in future studies.

Interestingly, Larsen et al. (2024) also identified the same pattern for chronic knee pain in individuals with facilitatory CPM profiles who had more positive self-reported function.[34] Though the study was cross-sectional and our study was post-intervention with follow-up, the convergence suggests that lower CPM has stratification value and is not maladaptive at any time point. With replication in a larger sample, this finding has the potential to inform individualized treatment planning (including discharge planning).[11]

ROC-optimized threshold values enhance the performance of CPM as a dynamic biomarker, highlighting how thresholding optimization may more effectively represent the potential to recover than binary divisions based on fixed cutoffs. The open publication of this analysis demonstrates our overall approach to the assessment of predictive thresholds and cutoff optimization techniques. Early results indicated the need for larger-scale validation studies to confirm and optimize post-mobilization CPM subgrouping thresholds for clinical use. Our findings on post-treatment CPM subgrouping as a dynamic biomarker offer promising new avenues beyond manual therapy for chronic pain. Although protocol adaptations to accommodate diverse clinical settings (e.g., perioperative timing for surgery or pharmacokinetics for drug trials) can be anticipated, standardized and validated dynamic CPM testing performed immediately before and after post-intervention could become routine. Most importantly, our approach uses maximally discriminant ROC/AUC-based cut-off points to stratify responders in an objective fashion, this may have research implications in surgical and pharmacologic scenarios with modification of the protocol. For example, perioperative assessment of acute central pain modulation changes may be used to guide individualized postoperative rehabilitation, and CPM measurement before and after drug intake may permit the on-time prediction of drug effects. Synergizing these pharmacologic and surgical dynamic CPM models with rehabilitation protocols can significantly enhance early patient stratification and individualized treatment pathways along the pain management continuum, positioning CPM as a real-time *dynamic* biomarker for precision pain medicine.

### Limitations

The methodological advantages of the current study include the implementation of a standardized spinal mobilization technique by an experienced therapist, blinded outcome measurement, application of more than one valid outcome measure, and a clear description of subgroup and continuous analyses. These design features add methodological strength to exploratory studies.

However, this study had several limitations. The small sample size restricts statistical power, compromises estimation precision, and limits the generalizability of the findings. Due to the exploratory nature of this study and the relatively small sample size, no corrections for multiple comparisons were applied; therefore, findings should be interpreted with caution. This trial recruited only adults with mild chronic mechanical neck pain, limiting its generalizability to other populations. The trial assessed only one post-intervention measure of CPM, which limited our understanding of the temporal dynamics of changes in CPM. Although bias reduction by blinding and protocol adherence minimized bias, the post hoc subgroup and ROC-derived cut points were exploratory rather than conclusive. The study was retrospectively registered to align with the shifting mechanistic hypotheses after data collection. Finally, selection bias was possible because of the recruitment procedure. Larger-scale prospective studies are required to validate post-mobilization CPM subgrouping, optimize threshold selection, and support the clinical application of this biomarker for personalized rehabilitation.

## CONCLUSION

These pilot findings illustrate the potential of post-spinal mobilization CPM subgrouping as a biomarker/phenotype measure in musculoskeletal rehabilitation. The integration of optimal CPM cutoffs into this approach strengthens early discharge prediction and further refines it as a mechanism-based clinical tool to abbreviate rehabilitation courses and foster efficient, individualized recovery. Validation in larger, more heterogeneous populations is needed to confirm these promising results and further advance post-SM CPM subgrouping as potential biomarker candidates for precision musculoskeletal rehabilitation.

## DATA AVAILABILITY

The de-identified dataset supporting the findings of this study will be publicly available upon publication.

## Author Contributions

EY, MW, JC, SY, and TS contributed to the design and implementation of the research; EY, SF, JAS, and AH analyzed the results and wrote the manuscript. EY conceived, and MW, SY, and JC supervised the project.

All the authors have no conflicts of interest to declare.

## Acknowledgement

The authors would like to thank Sherri Weiser, PhD, Shira Weiner, PT, PhD, Ali Sheikhzadeh, PhD for their mentorship with the NIOSH grant/subaward acquisition, and to Jason K Grimes, PT, DPT, PhD for his contribution with some data collection.

Post-Spinal Mobilization Conditioned Pain Modulation Stratifies Recovery Trajectories in Chronic Neck Pain: Toward a Mechanism-Informed Biomarker for Precision Rehabilitation

